# What are effective strategy to constrain COVID-19 pandemic crisis? lessons learned from a comparative policy analysis between Italian regions to cope with next pandemic impact

**DOI:** 10.1101/2022.05.15.22275107

**Authors:** Mario Coccia, Igor Benati

## Abstract

The pandemic of Coronavirus Disease 2019 (COVID-19) and its variants is rapidly spreading all over the world, generating a high number of infections, deaths and negative impact on socioeconomic system of countries. As vaccines and appropriate drugs for treatment of the COVID-19 can reduce the effectiveness in the presence of variants and/or new viral agents, one of the questions in social studies of medicine is effective public policy responses to reduce the impact of COVID-19 global pandemic and similar infectious diseases on health of people and on economies. This study analyzes public policy responses to the pandemic crisis across Italian regions that were the first areas to experience a rapid increase in confirmed cases and deaths of COVID-19. The analysis of regional strategies, from January to July 2020, reveals differences in public policy responses to delay and reduce the height of epidemic peak and to afford health-care systems more time to expand and respond to this new emergency. Veneto Region in North-East Italy has managed health policy responses with: a) a timely and widespread testing of individuals, b) units of epidemiological investigation for tracing all contacts of infected people in an effective contact tracing system. This public policy response has reduced total deaths and the final size of COVID-19 pandemic on health of people. Other regions have done public interventions without a clear strategy and goals to cope with diffusion of COVID-19 and as a consequence, they have had a higher negative impact on public health. Lesson learned can be important to design an effective public policy that can be generalized in different regional and national systems to prevent and/or reduce future epidemics or pandemics similar to the COVID-19.

## 1. Introduction

A new coronavirus (SARS-CoV-2) is the cause of a disease, Coronavirus Disease 2019 (COVID-19), which is rapidly spreading all over the world, generating a high number of infections and deaths from January 2020 (Coccia, 2022). COVID-19 produces minor symptoms in most people, but it is also the cause of severe respiratory disorders and death of many individuals worldwide (Coccia, 2020). Transmission dynamics of COVID-19 is due to manifold factors, such as density of people, air pollution commercial activity, unsustainable environments, socioeconomic factors, climate (cold temperature and humidity), new variants of SARS-CoV-2, health expenditures of countries, nonpharmacutical interventions, etc. (Bontempi and Coccia, 2021; Bontempi et al., 2021; Coccia, 2018, 2020a, 2021, 2021a, 2021b, 2022a, Coccia and Bellitto, 2018). The Coronavirus infection is an on-going global health problem that is also generating a socioeconomic crisis and negative world economic outlook projections (Sadat et al., 2020). In this context, national and regional public policies play a main role to cope with the COVID-19 pandemic crisis when effective drugs and treatments lack. Nicoll and Coulombier (2009, p. 3ff) argue that strategies to cope with pandemic can be based on:

- *Containment* measures have the goal to stop as many transmissions as possible and eventually the outbreak ‘burns out’. The prevention of an infectious disease is given by case-finding (detecting imported infections and first-generation transmissions) and taking actions to prevent their turning into chains of transmission and outbreaks, through vigorous contact tracing, treatment, quarantine of contacts and general lockdown of people (cf., Coccia, 2021c, 2022b)
- *Mitigation* measures decrease the impact of a pandemic by reducing number of people infected, reducing transmission, ensuring healthcare for those who may be infected, maximizing care for those with disease and protecting the most vulnerable, such as social distancing, school closures, workplace distancing, to avoid crowded places, etc.

Mitigation policies are highly recommended by World Health Organization when containment measures are no more possible in the presence of a high number of infected people. In the presence of COVID 19 crisis, many countries initially propose a containment policy (quarantine and general lockdown) and subsequently mitigation measures, such as social distancing, reduction of the interval between symptom onset and isolation, disinfection of buildings, masks for people in outdoor and indoor environment, travel restrictions, etc. (Kucharski et al., 2020; Nussbaumer-Streit et al, 2020; Walensky and Del Rio, 2020).

In this context, the study has two goals. The first is to analyze the differences of strategies of Italian regions to constrain the COVID-19 pandemic. The second goal of the paper is to determine what type of regional strategies and why is more effective to cope with the spread of the COVID-19 when drugs and effective treatment lack. Results of the study here can explain weaknesses and strengths of different regional responses in order to suggest a more effective regional and/or national strategy of public policy to guide policymakers to cope with future epidemics similar to the COVID-19.

## 2. Methodology

This study focuses on case study of Italy and specific regions of North Italy. The methodology of this study is based on qualitative and quantitative techniques. Qualitative techniques here analyze public policy at national and regional level in Italy using information from regional orders and decrees. A narrative approach is applied to analyze national and regional policies to cope with COVID-19 with two perspectives (cf., Sebeok, 1988): *a) temporal perspective* analyzes events of strategies anti-pandemic crisis over time (*chronos*) b) *opportunity perspective* analyzes conditions for which an event has occurred (*kairos)*. The interaction of these perspectives (time and opportunity) generates a critical explanation of relationships underlying public policies to cope with the COVID-19 pandemic crisis. In particular, this study performs a comparative analysis of public policies anti-COVID-19 in Veneto and Piedmont regions, which have comparable demographic and socioeconomic characteristics (Table 1).

**Table 1.** Demographic and socioeconomic characteristics of Veneto and Piedmont in Italy

| Region | Italy | Population,<br>2019<sup>a</sup> | Population<br>Density,<br>inhabitants/km<sup>2</sup><br>2019<sup>a</sup> | Gross Domestic<br>product per<br>capita (Euro),<br>2017<sup>a</sup> | Human<br>Development<br>Index, 2017<sup>b</sup> | Regional<br>Health<br>Expenditure, 2018<br>(millions of euro)<sup>c</sup> |
| --- | --- | --- | --- | --- | --- | --- |
| Veneto | Northeast | 4 909 013 | ~268 | €33,100 | 0.890 | 8 441,7 |
| Piedmont | Northwest | 4 356 406 | ~172 | €30,300 | 0.887 | 9 400,3 |

*Sources:* a) ISTAT (2020); b) UNDP (2020); c) Ministero dell'Economia e delle Finanze (2019)

In addition,

□ Veneto region, located in Northeast Italy, is one of the first regions, with Lombardy, to experience a rapid increase in the diffusion of COVID-19 but also in applying policy responses oriented to containment measures to cope with COVID-19 in society.
□ Piedmont region in Northwest Italy, developed an outbreak of COVID-19 later than Veneto, and is a comparable and representative region of other regions in North Italy that applied anti-pandemic measures with different effects.

Quantitative techniques here analyze the dynamics of COVID-19 related infected individuals and deaths in Veneto and Piedmont to determine the final impact of public policy responses in society (cf., Coccia, 2018a). Data from February to July 2020 by Ministero della Salute (2020) are used. In particular, number of confirmed cases and deaths are showed as total number and per million of people in June 2020 (the final phase of COVID-19 outbreak in Italy). Moreover, trends of daily variation of deaths and confirmed cases in Veneto and Piedmont (i.e., deaths/confirmed cases at *t* − deaths/confirmed cases at *t*-1) are represented as function of time and estimated with Ordinary Least Square method using a quadratic model. Estimated relationships of Veneto and Piedmont are also analyzed with an optimization approach to calculate the max number of deaths per million in the peak of COVID-19 pandemic crisis in these two regions under study. This analysis shows the final effect of pandemic of COVID-19 on public health in Veneto and Piedmont for a comparative evaluation of public policy responses to suggest effective public interventions to cope with future epidemics of the COVID-19 and similar viral agents

## 3. Results

### 3.1 National public policy to cope with COVID-19 in Italy: from containment to mitigation measures

The World Health Organization (WHO) claimed that COVID-9 has the level of a pandemic on 30 January 2020. Italy reacted immediately by establishing restrictive health measures for ship and air traffic on February 1^st^, 2020. On February 20, 2020, the first Italian case was diagnosed in Lodi, a small city bordering large urban conurbation of Milan in Lombardy region (Northwest Italy). The day after the number of confirmed cases rose rapidly to 36 infected people and subsequently the diffusion of COVID-19 in specific cities of Northern Italy accelerates. Italy initially applied a local containment strategy on some cities of the country. Italian government imposed on 23 February 2020 the so-called Red Zones, in areas with coronavirus outbreaks, based on lockdown of all commercial activities, restrictions to people’s freedom and mobility, quarantine of confirmed cases and appropriate home isolation of inter-related people (cf., Wells et al., 2020). These containment measures were initially applied only in two municipalities, Vò Euganeo in Veneto and Codogno in Lombardy region. Subsequently, these containment measures were extended to 11 municipalities, and general mitigation measures to the territory of six regions: Emilia Romagna, Friuli Venezia Giulia, Lombardy, Veneto, Piedmont and Liguria in North Italy. On March 10th, 2020, by decree of the Premier, Italy implemented the extension of containment measures throughout the country by total quarantine of people and lockdown of activities, except essential services such as groceries, postal offices, etc.: *Phase 1 of containment* from 10 March 2020 to cope with COVID-19 pandemic crisis in Italy (Governo Italiano, 2020). In this way, the general aim of Italian government was directed to reduce the height of epidemic peak, affording health-care systems more time to expand and respond to emergency and, as a result reducing the final size of COVID-19 epidemic (cf., Fong et al., 2020; Peak et al., 2017; Prem et al., 2020). In the context of containment, non-pharmaceutical measures in Italy included physical distancing, school closures, no sportive and cultural events, workplace distancing, masks wearing, etc. similarly to measures applied for COVID-19 outbreak in Wuhan, China (cf., Peak et al., 2017). In short, containment and mitigation actions can overlap and are often implemented concurrently, such as in Hubei (China) and, more recently, in some European countries, namely France, Italy and Spain (OECD, 2020). The national strategy of containment was immediately implemented by all twenty Italian regions but with different modes within regional health systems, producing different effects on public health and society. In particular, Veneto, in addition to the implementation of national measures, adopted a *specific containment strategy associated with proactive testing and in-depth tracing of infected cases*. The study of Veneto strategy and its results in controlling and reducing infections of the SARS-CoV-2 could offer some hints to discover the most effective public policy to cope with the COVID-19 pandemic crisis in the short and long run.

### 3.2 Comparative analysis of regional policy responses to constrain COVID-19 pandemic crisis: Veneto vs. Piedmont in Northern Italy

The public policy to constrain the COVID-19 pandemic crisis designed by government in Italy was aimed at reducing the spread of infectious diseases for delaying and lowering the height of epidemic peak. National public policy, however, was implemented in different contexts of Italian regions. Many regions applied only national guidelines of containment in a strict way, but other regions−using their administrative autonomy−applied additional measures that probably empowered those of the central government. A comparative analysis between regional policy of Veneto and Piedmont (a comparable region with the Veneto for its socioeconomic characteristics) provides interesting results.

In Veneto, on February 21, 2020, the town of Vò Euganeo^1^ had a COVID-19 outbreak and containment measures with Red Zone were applied to stop the diffusion of novel coronavirus with 14 days of full lock-down of this city, quarantine and a ban on population movement from 23^rd^ February to 8^th^ March 2020 (Lavezzo et al., 2020).

In Piedmont, the first confirmed case of COVID-19 was detected on February 22, 2020, a 40-year-old man having a contact with infected people from Lombardy (the biggest COVID-19 outbreak in Italy). The day after, the regional government adopted national guidelines of the Health Ministry of Italy, stating the suspension of all events and any type of aggregation in a public or private places, the closure of children’s education services, schools, universities, museums and other places of culture. In addition, regional government in Piedmont established a legal obligation to people from urban areas with high epidemiological risk to inform healthcare system for the adoption of fiduciary home isolation with active surveillance. Initially, in Piedmont there was not any local outbreak, but infected individuals were mainly in provinces bordering Lombardy region (e.g., Asti and Alessandria). On March 8th, 2020, five out of eight provinces in Piedmont were declared Red Zones, with a full lockdown, because of growing number of the confirmed cases of COVID-19 (Regione Piemonte, 2020).

In Veneto, after the declaration of Red Zone in the town of Vò Euganeo, all population of this town was tested twice for the presence of the novel coronavirus using nasopharyngeal swabs. The fieldwork, led by University of Padova, clarified some key aspects of the transmission dynamics of the SARS-CoV-2 (e.g., the active role of asymptomatic people) and triggered a learning process for supporting the strategy of containment of Veneto.

As of 29 February 2020, 3% of the population was found positive to the novel coronavirus and many people were asymptomatic in this town ^2^. People with positive tests (i.e., having the COVID-19) were placed in home isolation. By March 23^rd^, 2020, the spread of the COVID-19 had been stopped and no new cases could be identified in the town of Vò Euganeo in Veneto. This initial medical trial suggested that a widespread testing of people with no symptoms, associated with isolation of infected individuals, was a critical strategy to control the spread of COVID-19 pandemic. On the basis of this experience, the University of Padova proposed to the Region of Veneto to adopt a proactive strategy for COVID-19 through the use of swabs extensively across the population of this region. The proactive strategy of swabs in Veneto was published with the Regional Decree No.344 of Veneto on 17^th^ February 2020 (Regione Veneto, 2020), before the containment measures of Italy started on 10^th^ March 2020. The strategy anti-COVID-19 by Veneto had the following structure:

° *General goal*: “to stop the transmission chain of the virus responsible for COVID-19” in the general population through identification of “positive” paucisintomatic and asymptomatic subjects, enlarging the home isolation around the “positive” case (Regione Veneto, 2020a).
° *Specific sub-objectives* given by:

- identify all possible suspicious, probable and confirmed cases of people with COVID-19
- apply for all people having contacts with infected individuals, the measures of quarantine and fiduciary home isolation
- re-organize the activity of the Regional Prevention Departments to support of the COVID-19 pandemic crisis
- screening of all Regional Health System employees and inter-related subjects to enhance public safety of these subjects and patients
- find all positives cases of COVID-19 in employees of Essential Services.

The strategy of Veneto, based on general and specific goals, was unique among all Italian regions. It was implemented by the Prevention Departments of the Veneto with the collaboration of the University Hospital of Padova and the Italian Red Cross Committee, through the coordination of the Prevention, Food Safety and Veterinary Directorate of the Veneto.

The implementation of this anti-COVID-19 strategy was based on four main elements:

° *Target population*.
  a. individuals potentially connected to a cluster or exposed to infected people (e.g., family, work, or social /occasional contacts of suspicious or confirmed cases) that have been and may have had a link with a confirmed or probable case of COVID-19. The investigation has to be focused on previous 48 hours from initial symptoms to the diagnosis and home isolation;
  b. healthcare employees and essential services employees such as shopping malls employees, firefighters and police officers.
° *Swabs*. The requests for swabs have been done by the General Physicians and Pediatricians of Territorial services transmitting appropriate information to Territorial Operational Unit (COT) of the Local Health Unit, or by other medical specialists that had to report them to Public Health Hygiene Service (SISP) or COT according to local organization. The COT and the SISP had to communicate the list of people to test to the lab identified for the execution of swabs.
° Guidelines for subjects. The subject with symptoms had to stay at home until the swab is done and result is received. Home isolation was compulsory for people having a link with confirmed or suspect case of COVID-19. Asymptomatic people could have swabs in specialized health structures. In this context, a main element of the strategy was the epidemiological investigation with increasing concentric cycles of inquiry starting from the individual, possible contacts in family and at workplace at high risk, to social and occasional contacts at low risk. In the presence of a confirmed case, all their contacts had to stay in home isolation. Confirmed cases were subjected to an “active surveillance” in isolation (with daily follow-up telephone call or a telemedicine Service). In fact, a rapid contact tracing is basic within epicenter to limit human-to-human transmission outside of outbreak areas, also applying appropriate isolation of cases (Wells et al., 2020).

To implement its strategy, the Veneto used 13 laboratories for testing the SARS-CoV-2. Medical technology had high performance in terms of the number and speed of tests because of large stocks of reagents and equipment for lab analysis with open technology, i.e., capable of using reagents of different brands (Primatreviso, 2020). Swabs could be done at hospital, at home or in the car. In short, Veneto applied a proactive strategy of swabs to symptomatic and asymptomatic people, with an in-depth epidemiological inquiry for detecting all contacts and subsequently home isolation of confirmed cases and suspected people.

Piedmont, by contrast, applied containment measures in accordance with national guidelines of Italy, with a strategy of confirmative swabs for infected and suspected people. In this region, swabs (nasopharyngeal and oropharyngeal) were used only to confirm symptomatic suspect cases and to certify healed individuals; there was not any testing for searching asymptomatic people of COVID-19. Kucharski et al. (2020) claim that the isolation of cases and contact tracing can be less effective for COVID-19 because infectiousness starts before the onset of symptoms (cf., Peak et al., 2017). Hellewell et al. (2020) show that the probability of control decreases with long delays from symptom onset to isolation that increase transmission before symptoms. Piedmont also made swabs both in hospitals, at home and in the car but general physicians were authorized from May 11, in the maturity phase of the COVID-19 outbreak and about three months later than Veneto. In addition, at the beginning of the COVID-19 pandemic, in Piedmont, only two laboratories were active for tests with the capacity of making about 400 swabs per day, whereas in Veneto were 13 active labs from 17^th^ February 2020 onwards. Later, the capability of test in Piedmont had been greatly increased, with the opening of new labs but the COVID-19 had already generated issues for public health of this region. Although, the increased potential to perform diagnostic tests for COVID-19, Piedmont did not apply neither an active case-tracking strategy by swabs, nor the activation of in-depth epidemiological inquiries to detect symptomatic and asymptomatic cases of people that have had social and occasional relationships with infected people.

Both regions have also reorganized their healthcare facilities for COVID-19. Veneto and Piedmont have set up specialized hospitals for the treatment of patients with COVID-19 and increased the number of Intensive Care Unit (ICU) beds. Veneto increased ICU beds from 494 to 825 on April 29^th, 2020^; Piedmont, from 327 to 827 ICU beds at the same data. Finally, about laboratories for testing the SARS-CoV-2, Veneto had the operational capacity of 13 laboratories throughout the pandemic crisis, whereas Piedmont had only two laboratories at the beginning of the crisis, subsequently the number increased to a total amount of 20 laboratories in the phase of maturity of the cycle of life of COVID-19.

The comparison between Veneto and Piedmont suggests some vital policy implications (Table 2). The two regions under study here applied, in general, containment measures, based on national guidelines of Italy. However, Veneto integrated containment measures with an initiative-taking testing and in-depth epidemiological inquiry of infected cases creating a fruitful combination to stop as many transmissions as possible and local COVID-19 outbreaks. In short, Veneto, using the experience and learning of effective procedures applied in the initial outbreak of the town of Vò Euganeo, implemented a proactive testing and tracing of people, unlike Piedmont.

**Table 2.**
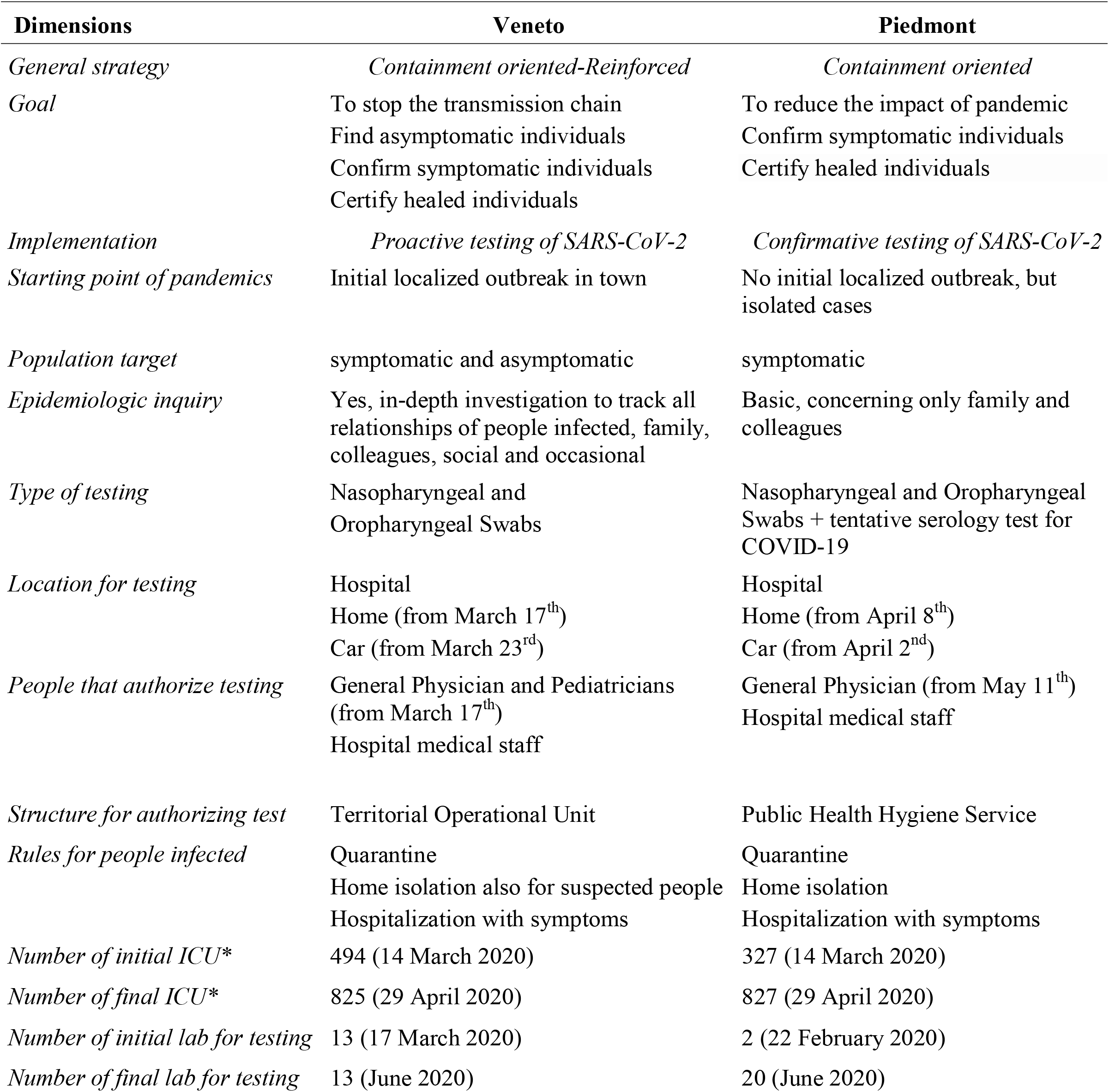
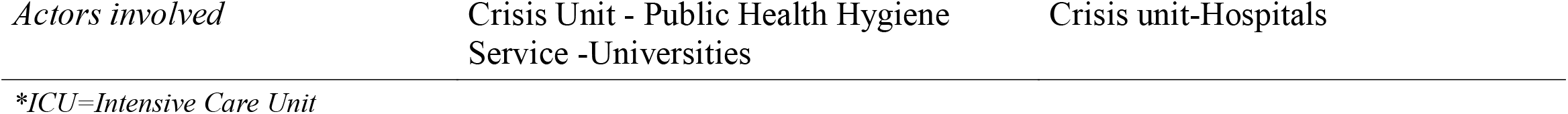
Comparative policy responses in Veneto and Piedmont Region

### 3.3 The dynamics of the COVID-19 in Veneto and. Piedmont

The study of the comparative dynamics of COVID 19, deaths and swabs in Veneto and Piedmont shows some relevant results of the different public policies implemented by these regions from phase 1 of containment (about February to May 2020) to phase 2 of mitigation measures (from June 2020 onwards).

Figure 1 shows that at beginning of the outbreak the number of daily confirmed case in Veneto was higher than Piedmont. Subsequently, after the proactive strategy of testing and tracing to cope with COVID-19, the dynamics of infected cases is always lower than Piedmont with reduced daily variation. Figure 2 shows that trend of swabs in Veneto is always higher than Piedmont. In addition, figure 2 reveals that at beginning of COVID-19 outbreak Veneto and Piedmont had a rather similar trends of deaths per million people. Since mid-March, Veneto has recorded a systematically number of total deaths lower than Piedmont. This evidence, using confirmed cases, number of swabs and total deaths, suggests that public policy applied by Veneto has been more effective to cope the COVID-19, reducing the final size of COVID-19 epidemic on public health.

**Figure 1.**
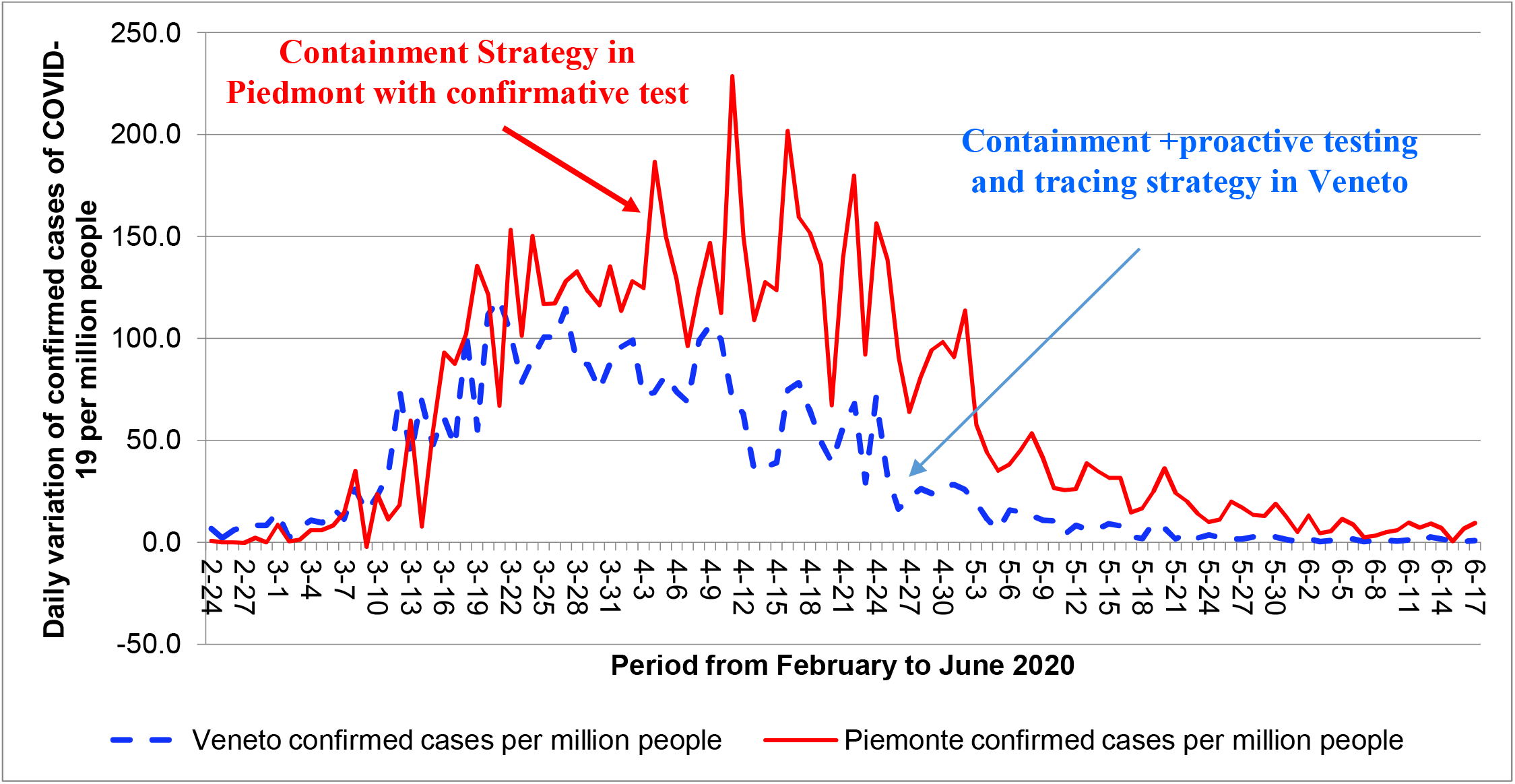
Trends of daily confirmed cases of COVID-19 in Veneto e Piedmont

**Figure 2.**
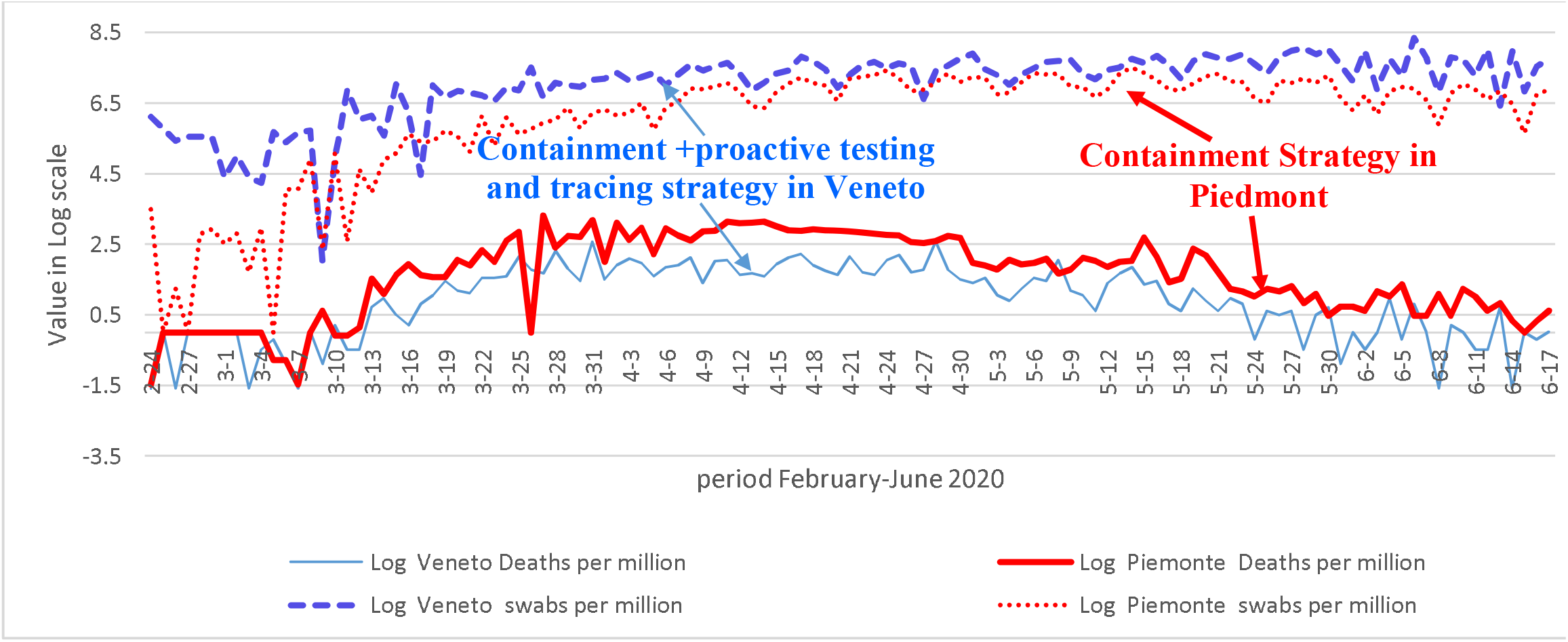
Trends of deaths and swabs in Veneto and Piedmont

In order to confirm this result, Figure 3 shows the estimated relationships (quadratic models) of deaths per million over time (cf., Table 3). The models show a high goodness of fit with a high coefficient of determination (R^2^), suggesting that approximately 60% of the observed variation of COVID-19 related deaths can be explained by independent variable of time. The application of optimization technique on these equations shows that the peak of deaths is about at 56^th^ days from 24^th^ February 2020 both in Piedmont and Veneto.

**Table 3.** Estimated relationship of total number of deaths in Piedmont and Vento (quadratic model)

| Estimated relationships (quadratic model) | $R^2$ |
| --- | --- |
| Deaths per million people (Piedmont)= $-0.0056t^2 + 0.6248t - 3.4004$ | 0.5951 |
| Deaths per million people (Veneto)= $-0.0023t^2 + 0.2599t - 1.2224$ | 0.6061 |

Note: Explanatory variable is time= $t$

**Figure 3.**
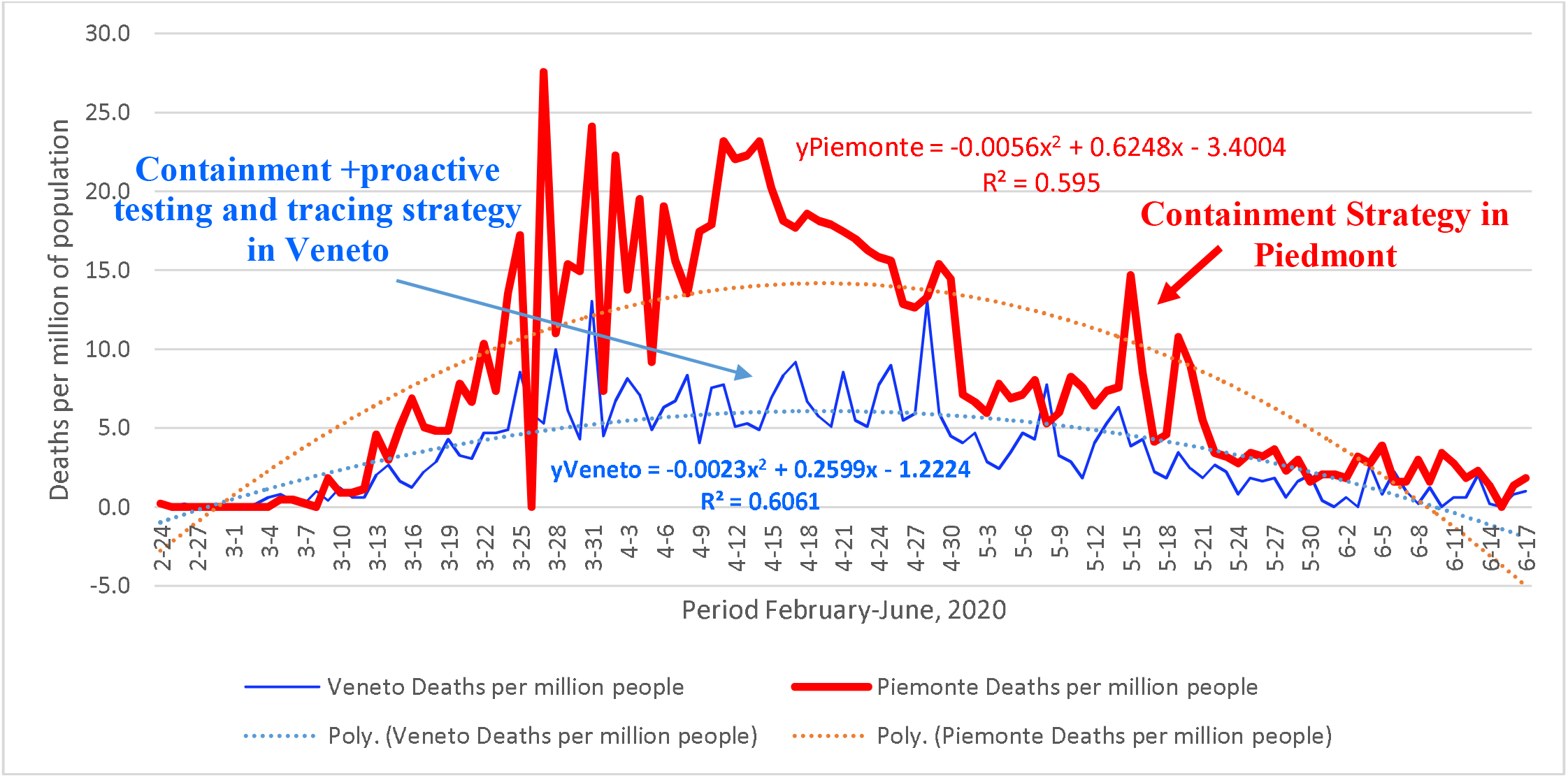
Trends and estimated relationship of deaths over time in Veneto and Piedmont regions

However, a main difference is given by max number of total deaths of these estimated relationships indicated in Table 4: Veneto has a number of total deaths equal to 6.12 per million people, a value that is less than half compared to Piedmont (given by 14.02 per million people). To put it differently, Piedmont has had +129.0% of total deaths in the peak of the COVID-19 than Veneto! *Results in table 3 show that Veneto, with a containment approach reinforced with a high number of swabs to detect symptomatic and asymptomatic people and in-depth epidemiological inquiries has probably reduced total number of deaths of about the half compared to Piedmont*.

**Table 4.** *Max* number of total deaths per million of people in estimated relationship of Veneto and Piedmont

|  | **Veneto** | **Piedmont** |
| --- | --- | --- |
| <u>Max number of total deaths per million of people</u> | 6.12 | 14.02 |
| <u>$\Delta\%$ of Piedmont vs. Veneto</u> | -- | +129.09% |

Table 5 confirms the effectiveness of regional policy responses by Veneto to the COVID-19 pandemic crisis. This public policy can suggest an efficient strategy to cope with future epidemics similar to the COVID-19.

**Table 5.**
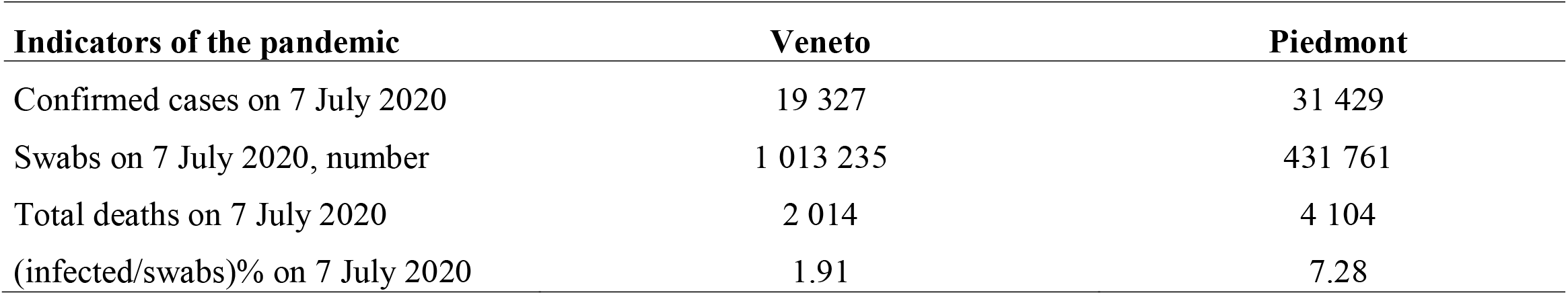
Final size of COVID-19 pandemic crisis on public health in Veneto and Piedmont Region

| **Indicators of the pandemic** | **Veneto** | **Piedmont** |
| --- | --- | --- |
| Confirmed cases on 7 July 2020 | 19 327 | 31 429 |
| Swabs on 7 July 2020, number | 1 013 235 | 431 761 |
| Total deaths on 7 July 2020 | 2 014 | 4 104 |
| (infected/swabs)% on 7 July 2020 | 1.91 | 7.28 |

## 4. Discussions and conclusions

The effective management of the COVID-19 global pandemic crisis and its effects on socioeconomic systems require more than the actions of healthcare setting alone. In fact, in the presence of COVID-19 outbreaks, it is crucial to understand national and regional policy responses to the pandemic crisis for designing strategies to stop and/or reduce accelerated diffusion of viral agents (Coccia, 2020). This study focuses on Italy that is case study particularly interesting because national strategy was oriented to a containment policy, and it was implemented and/or integrated by regions with different modes and timing. This variety of regional applications of national orders, coupled with regional health system, has produced different public interventions and, as a consequence, different impacts on public health and society (cf., Benati and Coccia, 2019)

This study shows that regions under study adopted different strategies of testing and of epidemiological inquiries (fig. 4). On one hand, Veneto adopted a proactive testing strategy for detecting suspicious, probable and confirmed cases, moreover, the regional Public Health Hygiene Service carried out an in-depth epidemiological investigation by specific units in the Department of Prevention able to guarantee swabs to paucisintomatic or momentarily asymptomatic subjects that can be potentially linked to a cluster (cf., DGR, 2020). The outcome of this strategy was the reduction of the period from symptom onset to isolation that reduces human-to human transmission before symptoms, and as a consequence the effects on public health in terms of a lower number of deaths (cf., Hellewell et al., 2020; Kucharski et al., 2020).

**Figure 4.**
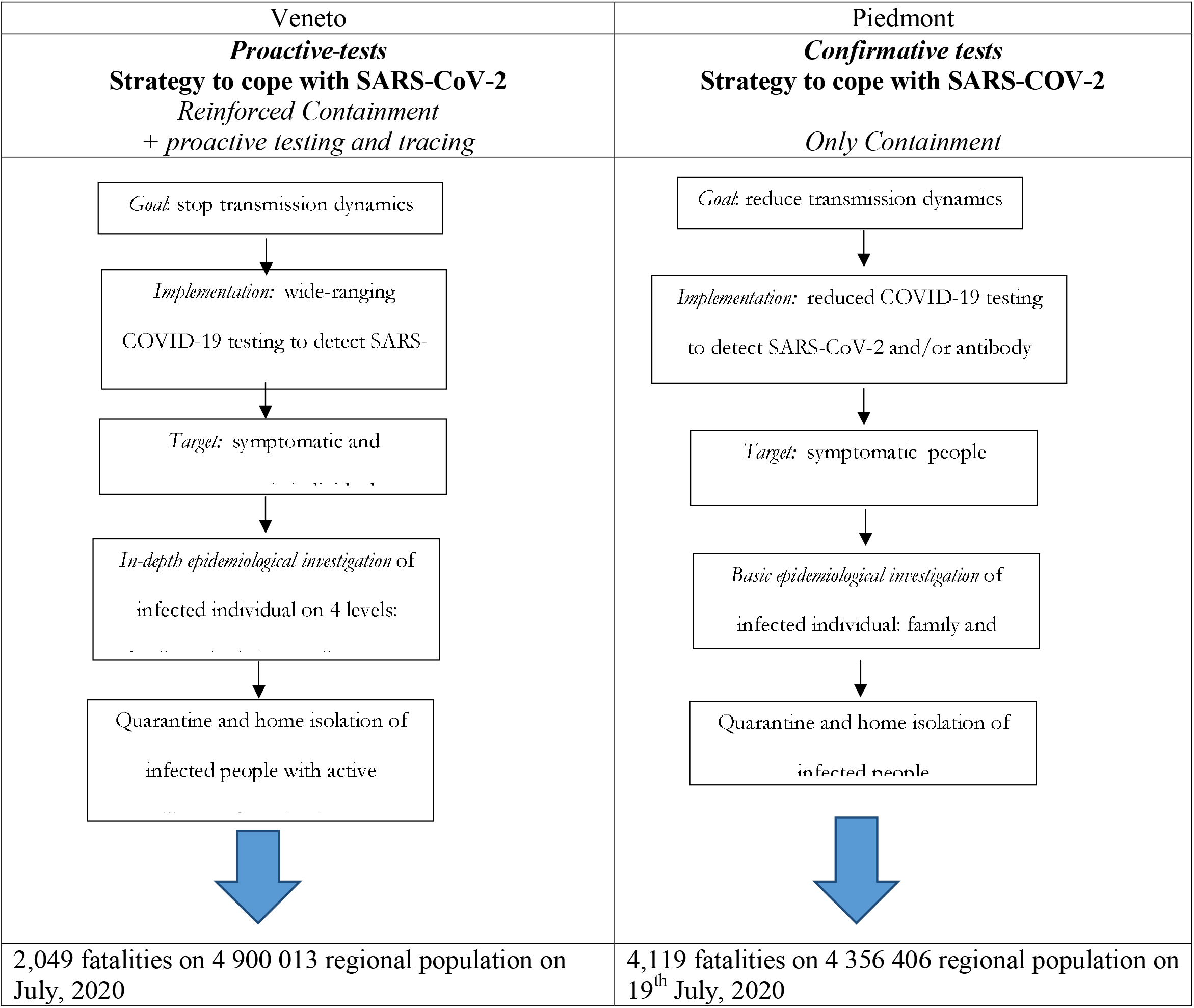
Different policy responses to the COVID-19 pandemic crisis

On the other hand, Piedmont did not design complementary measures based on proactive testing strategy of people and systematic epidemiological inquiries of all contacts of confirmed cases, like Veneto, and applied only the general indications of national government. In particular, Piedmont has limited the epidemiological investigation to detect possible links in the family and at work of confirmed cases. The outcome is a longer delay from symptom onset to isolation that increase transmission of COVID-19 with a high negative impact on public health in terms of number of total deaths (cf., Hellewell et al., 2020).

In the presence of a COVID-19 global pandemic crisis given by an unexpected complex problem that threats cities, regions and nations (Bundy et al., 2017), this study suggests the opportunity to adopt a policy response based on initially on containment measures, integrated with a proactive and widespread tests of people and in-depth epidemiological investigations of all contacts, and subsequently mitigation measures, applying as far as possible a rapid decision making in order to reduce socioeconomic problems in the short, medium and long run (Klein, 1993). To put it differently, an effective public policy response to the COVID-19 should not only implement measures of lockdown, quarantine and social distancing measures (containment and mitigation measures) but it must be complemented by a proactive testing strategy of people and in-depth epidemiological inquiries on a large scale.Lessons learned are that effective strategies to cope with COVID-19 must have general and specific goals to cope with the spread of infectious diseases within a general regional system in which policy networks, institutions, scientific and technical expertise, and processes of learning interact in regional system (Coccia, 2018b, 2019, 2021d, 2021e, 2022c, 2022d). In fact, Lasswell (1956) argues that effective public policy requires the understanding of the dynamics of processes, actors, and interactions that shape policy decisions in response to the crisis, such as for COVID-19 pandemic crisis. In addition, the effect of crises can be worsened by weak institutions, inefficiencies and delays in policy responses, both at the regional and national level. Some vital elements that have supported the efficiency and effectiveness of policy responses of Veneto can be generalized as follows:

□ Leveraging *operational levels* based on medical personnel, epidemiologists, biologists, emergency managers, and other professionals coping with the pandemic’s immediate threat. This level is underpinned in a *strategy* in which political leaders carry responsibility, make strategic decisions, provide funds and support coordination and collaboration among different subjects and units (Boin and ’t Hart 2010).
□ *Network of innovators* with a great variety of expertise in different fields plays a vital role to support policy decisions and their implementation (Jenkins-Smith et al., 2018). For example, the vital role of the University of Padova and some leading professors.
□ In the presence of crisis and high uncertainty, *the demand for scientific and technical expertise* increases for guiding decision making of policymakers, such that decisions are based on high scientific expertise that serve the public good, rather than specific interests (Cairney 2016). In Veneto, but mainly in Italy, many decisions to cope with COVID-19 pandemic crisis are supported by scholars of academia and public research labs that serve to legitimize and justify government responses to consequential problems of the COVID-19 pandemic crisis. Hence, in the presence of uncertain problems, scholars play a vital role in supporting public policy responses and are part of the rationale of governments’ responses (Webley et al., 2020).
□ A main element to support policy responses to cope with COVID-19 pandemic crisis is *learning* that plays a critical role in the ability of understanding complex policy issues and improving public policy. Learning of Veneto with the initial outbreak of the town Vò Euganeo has supported innovative and effective policy responses (cf., Crow et al. 2018).
□ Finally, the implementation of the public policy is not self-enacting. Every aspect of implementation shapes how public policy takes place “on the ground”—from how administrators interpret policy directives to the way front-line personnel operationalize them, etc. (Weible et al., 2020). Policy responses require collaboration across different structures and organizational cultures, such as academic and administrative institutions (Hupe, 2013).

Overall, then, success or failure of policy responses to the COVID-19 pandemic crisis depend on manifold factors concerning human resources, institutions and organization. This study shows that a regional public policy of containment and mitigation measures, reinforced with a proactive strategy based on proactive testing of people and in-depth epidemiological inquiries on a large scale, can reduce total deaths and the final impact of COVID-19 and similar epidemics on public health. However, there are several challenges to such studies, particularly in real time. Sources may be incomplete, or only capture certain aspects of the on-going dynamics of public policies. In addition, public policies to constrain the socioeconomic and health effects of the COVID-19 pandemic crisis also include elements of bounded rationality of decision makers, aimed to a behavior of satisficing to cope with consequential environmental threats in the presence of uncertainty and highly restricted time for decision making (Gigerenzer and Selten, 2002). Hence, these conclusions are of course tentative in the presence of complex problems generated by COVID-19 global pandemic crisis and its transmission dynamics. To conclude, there is need for much more research into the relations between pandemic crisis of novel viral agents and effects of public policy on human health and society.

## Data Availability

All data produced in the present study are available upon reasonable request to the authors

## Footnotes

1 Vò Euganeo is a town (3,297 inhabitants) not far from Padova (30 km). It became the center of the COVID-19 pandemic in Veneto when two people were found positive on 21 February 2020. The next day, one of them, a 78-year-old man, died, the first COVID-19 death in Italy. On 24^th^ February 2020 an “iron-clad sanitary cordon” was created around the town and a testing program for all of its residents initiated driven by University of Padova.

2 An asymptomatic carrier is an infected individual with a pathogen that does not show symptoms and can transmit it to others, developing symptoms in later stages of the infectious disease.

